# A mobile app for chronic disease self-management for individuals with low health literacy: A multisite randomized controlled clinical trial

**DOI:** 10.1101/2023.04.01.23288020

**Authors:** Raymond L Ownby, Drenna Waldrop, Rosemary Davenport, Michael Simonson, Joshua Caballero, Kamila Thomas-Purcell, Donrie Purcell, Victoria Ayala, Juan Gonzalez, Neil Patel, Kofi Kondwani

## Abstract

**Objective:** The purpose of this study was to evaluate the effects of a mobile app designed to improve chronic disease self-management in older adult patients with low health literacy and who had at least one chronic health condition, and to assess the impact of delivering information at different levels of reading difficulty.

**Methods:** A randomized controlled trial was completed at two sites. Individuals 40 years of age and older screened for low health literacy who had at least one chronic health condition were randomly assigned to a tailored information multimedia app with text at one of three grade levels. Four primary outcomes were assessed: patient activation, chronic disease self-efficacy, health-related quality of life, and medication adherence.

**Results:** All groups showed overall increases in activation, self-efficacy, and health-related quality of life, but no change in medication adherence. No between-group differences were observed.

**Conclusions:** The mobile app was effective in increasing participants’ levels of several psychosocial variables, but reading difficulty level was not significantly related to outcomes.

Registered at clinicaltrials.gov NCT02922439.

## Introduction

Health literacy, defined as a person’s ability to find, take in, and use health information to attain a desired health status,^1^ and is related to health status and health outcomes across a wide range of contexts and health conditions. The 2003 United States (US) National Assessment of Adult Literacy showed that more than 75 million Americans had only basic health literacy skills, indicating that 1 in 4 Americans have problems understanding information about healthcare.^2^ More recent studies of literacy and numeracy skills in the general population suggest that the situation has not changed.^3, 4^ Research also shows that the problem is not limited to the US, with similar findings in Canada, Europe and Africa.^5–7^

Further, health literacy is lower in persons from racial and ethnic minoritized groups as well as older adults^2, 8^ and may be an important factor in health disparities.^9, 10^ In the US, 24% of Black persons (9.5 million) and 41% of Hispanic persons (21 million) have below basic levels of health literacy.^2^ Members of minoritized groups have lower levels of health literacy and compelling evidence links race and ethnicity to disparities in health via health literacy.^11–18^ Members of racially minority\ized groups and older adults are also more frequently affected by chronic diseases such as cancer, high blood pressure, heart attack, stroke, diabetes, asthma, hepatitis, HIV infection, mental health disorders and many others. The twin burdens of chronic disease and low levels of health literacy thus fall disproportionately on those most in need – members of minorities and older adults, all of whom may experience one or more chronic conditions while not having the health literacy skills they need to cope.^9, 19^ Interventions to improve health literacy are thus clearly needed.

Providing information for patients in clinical settings on self-management of their health conditions may be a useful strategy in addressing low health literacy. In the traditional methods that use pamphlets or handouts supported by conversation in a brief clinical encounter, however, information is often not actually read or remembered,^20, 21^ and recommendations may not be implemented.^22, 23^

One strategy to increase the impact of such information on patient behavior is tailoring. Tailoring information, defined as using various methods to create individualized communications for patients,^24^ aims to reduce the burden of self-management on health consumers by giving them useful information that is relevant to their needs or concerns and that they can understand and use. Various information interventions have been developed to improve patients’ health literacy,^25–27^ including matching health education content to patient characteristics and tailoring health messaging to make it more directly relevant to patients. While these techniques have often been successful,^25, 28^ creating individually-tailored health information is labor intensive and thus may not be widely available.

One solution to the problem of giving patients the information they want and need in a form they can use has been the development of computer-based interventions to automate message tailoring.^29, 30^ Computer-based tailoring creates the possibility that high-quality individualized health information can be made available to those who need it. Information can be delivered to patients when they want it and it can target content they are interested in, a process of providing precision health information.^31^ Analogous to the processes underlying the precision medicine approach to the somatic treatment of diseases,^32^ computer-based tailoring can take a patient’s personal characteristics, including their expressed concerns or problems, and provide detailed information to help them understand their health conditions and develop self-management skills.

A potentially critical variable in the tailoring process is ensuring that content is appropriate to the patient’s level of health literacy. If information is not understood, even if tailored, it may not impact patients’ behavior. Studies show that while experts recommend that materials for patients be written at a 4^th^ or 6^th^ grade level and match patients’ level of health literacy,^33–35^ multiple studies show that most patient education materials are at much more difficult levels.^36–40^

In this project, we worked to address these issues by developing a tailored information app focused on chronic disease self-management (CDSM)^41^ with three versions: one with text at 8^th^ grade level, a second at 6^th^ grade, and a third at 3^rd^ grade level supported by audio narration.^42^ We chose CDSM because we believed it was a logical target for a health literacy intervention. In an approach that cuts across specific diseases, CDSM targets problems and skills needed to cope with issues such as fatigue, pain, stress, depression, sleep disturbance and treatment adherence. Studies show that in-person CDSM classes improve patients’ functioning and reduce healthcare utilization^41, 43–48^ but their availability is limited due to the lack of qualified personnel, cost, and accessibility.

Similarly, while interventions have been developed to improve health literacy^12, 49^ they are difficult to scale to levels needed to meet the challenge of low health literacy (millions of persons worldwide) due to their cost^30, 50, 51^ Effective interventions with the potential for wider dissemination at reasonable costs are urgently needed. For many of the problems that are the focus of CDSM, well-defined behavioral strategies exist for their management (e.g., cognitive behavioral therapy for sleep^52^ or mood problems^53^) holding the possibility that effective tailoring might help patients develop relevant knowledge and behavioral skills.

Assessments of CDSM programs have focused on a several outcomes, and in this project, we focused on four that we judged were most relevant to the effects of a CDSM-targeting intervention and that would allow us to compare our results with those of other researchers. *Patient activation*^54^ is defined as the extent to which they are actively involved in their healthcare, and has been related to a number of important health status variables, including emergency department visits, receiving breast cancer screening as well laboratory measures such as hemoglobin A1C (related to diabetes) and HDL (high density lipoprotein related to cardiovascular disease risk).^55^ Activation has also been associated with self-management behaviors^56^ as well as quality of life,^57^ and studies show that interventions that improve patient skills can increase activation.^58^ Finally, activation is related to self-management behaviors in older adults^59^ and changes in activation are related to changes in health outcomes^60^ and healthcare costs.^61^

*Self-efficacy,* or a person’s belief in his or her capacity to reach specific goals,^62^ is a key concept in understanding health behavior,^63^ especially in relation to patients’ health behavior^62^ and in CDSM.^43^ In-person and internet-delivered CDSM programs have a positive impact on self-efficacy^45, 46^ which in turn have been related to self-management.^64–66^ Another highly relevant potential outcome of improved CDSM skills is improved *health-related quality of life* (HRQOL). Self-reported health status has often been studies as an outcome in CDSM studies,^41, 45, 47^ and is clearly an important aspect of improved self-management skills. Finally, *medication adherence* is an essential aspect of self-management behavior. Poor medication adherence is common in patient populations with estimates of adherence ranging from 55^67^to 75%.^68^ Adherence has been linked to numerous health outcomes,^69^ increased healthcare costs,^70^ and studied as an outcome in other studies of CDSM.^47^

The objective of this study was to assess whether a mobile app for CDSM providing individually-tailored health information would have a positive impact on participants’ activation, self-efficacy, quality of life and medication adherence. It was also hypothesized that information presented at reading levels consistent with expert recommendations (3^rd^ to 6^th^ grade levels) would have a greater impact on these variables than information presented at an 8^th^ grade level.

## Method

Initial guidance on content for the app was drawn from a review of existing sources on CDSM supplemented by a qualitative study that explored older patients’ information needs.^71^ A multidisciplinary team comprising representatives from medicine, nursing, psychology, pharmacy, public health, and education was assembled to develop content. Team members with special expertise in developing culturally and ethnically appropriate education materials were included as well (KTP, KK).

The app was conceived as a series of topical modules that would consist of a series of screens. Based in principles of cognitive load theory-based instructional design,^72, 73^ each module was planned to include an orientation to its purpose, assessment of the participants’ current status by way of questions, general health information on each topic, individually-tailored content, and a summary. Self-test questions were included to help participants understand how well they learned the module’s contents. Information was presented as text on a series of screens, supplemented by pictures, graphics, and narrated animations consistent with the principles of multimedia learning.^74^

More extensive information about the development and testing of the app and its contents are available in a paper under review, available as a preprint.^75^ Modules with the same content were created at three levels of reading difficulty based on the Fry^76^ and Flesch Reading Ease^77, 78^ scores of the text they contained (3^rd^ grade, with text narrated; 6^th^ and 8^th^) using Health Literacy Advisor® (Bethesda MD: Health Literacy Innovations LLC), a software plugin working with Microsoft Word®.

### Participants

Participants were recruited from participants in previous studies, from local health clinics and medical practices, and by word of mouth. At the Atlanta site, a paid recruiter visited local churches where she could screen potential participants as well as give them information about participation. Information and race and ethnicity was collected as required by the U.S. National Institutes of Health for grant recipients.^79^ Race and ethnicity were self-reported by participants. Gender was also self-reported, with transgender participants considered as the gender of their chosen identity.

### Screening

Participants were initially screened to determine their potential eligibility using a brief interview that elicited medical history, medication use, and education. They were administered a short form of the Rapid Estimate of Adult Literacy in Medicine^80^ using a previously validated cut-off for health literacy at or below the 8^th^ grade level.^81^

### Inclusion and exclusion criteria

To be eligible to participate in the study, participants were required to be 40 years of age and older, have at least one chronic health condition for which they were currently treated, have an education level less than 16 (i.e., not be a college graduate), and score below the cut-off score on the short form of the REALM. The criterion of having less than a college education was derived from our findings in a previous study^31^ that no participants who had successfully completed a college degree had inadequate health literacy.

### Measures

Participants completed an extensive battery of measures as part of a baseline assessment, with self-report outcome measures administered immediately after completing the intervention and then three months later. Most self-report measures were administered by computer, with questions read aloud by the interviewing software to minimize the impact of participants’ reading skill on their ability to respond to questions. As the baseline visit, participant demographic information, level of education, and medical history were assessed in an individual interview. The medical history interview was based on the medical conditions comprising the Functional Comorbidity Scale^82^ but expanded to include additional health conditions common in older adults.^83^ Self-report measures were administered via audio computer-assisted self-interview software (Bethesda MD: Questionnaire Development System) that read all questions aloud to participants to keep effect of reading ability on participant responses to a minimum.

In order to provide a standardized assessment of participants’ reading skills, trained assessors individually administered the Woodcock-Johnson Psycho-Educational Battery^84^ Passage Comprehension subtest. This measure provides a grade equivalent score that helped characterize participants’ reading levels. The FLIGHT/VIDAS health literacy scale was used in this study because of its desirable psychometric characteristics^85^ that include a wide range of scores; other health literacy measures often have ceiling effects that reduce the range of observed scores that make them less useful in statistical analyses.^86^

The four outcomes we studied were assessed with widely-used measures of each construct. Activation was evaluated with the Patient Activation Measure.^54^ We used the ten-item version based on the short form that has been shown to have good validity and reliability.^87, 88^ Self-efficacy was evaluated with the Chronic Disease Self-Efficacy Scale,^89^ used in multiple studies of CDSM programs. It also has demonstrated reliability and validity.^89^ The Medical Outcomes Study, Short Form 36 (MOS SF36^90^) is one of the most widely used measures for understanding psychosocial functioning related to health. It has well-established reliability and validity for use with older adults with chronic health conditions.^91–93^

The Gonzalez-Lu questions were included as a measure of adherence because they have been validated against electronically-recorded medication adherence and their simplicity.^94, 95^ As the four questions were highly intercorrelated, we reduced them to single score using principal axis factor analysis and used resulting factor scores as a single measure. This approach avoids the limitations of simply summing a group of items^96^ and has good psychometric properties.^97^

### Analyses

Planned analyses assessed the study hypotheses that persons receiving the intervention would (1) show significant increases in measures of activation, chronic disease self-efficacy, health-related quality of life and medication adherence and (2) that persons receiving the information at 6^th^ and 3^rd^ grade levels (experimental conditions) would show greater change than those receiving the material at the 8^th^ grade level (control condition).

Preliminary review of data and descriptive measures were obtained using SPSS version 28 (Armonk NY: IBM). Mixed effects random intercept models were evaluated with the statistical program R, version 4.2.1.^98^ using the *lme4* package.^99^ All models included participant age, gender, race, and site of data collection as well as time and treatment group and their interaction. Tests of study hypotheses were completed after maximum likelihood estimation using Satterthwaite approximations of *p* values.^100^ Tests of the statistical significance of within- and between-group differences were obtained using the *emmeans* package using the Tukey correction for multiple comparisons.^101^

Based on evaluation of between-group differences at the two sites as well as theoretical considerations, model covariates were chosen to control for likely confounders as well as observed between-site differences. They were chosen based on considerations of likely confounding impact on outcome measures (age, gender, race, and education) and observed between-site differences in level of health literacy and level of multimorbidity (FLIGHT/VIDAS health literacy scale and number of health conditions). In addition, because of observed differences in participant characteristics at the two sites, site itself was included as a covariate.

### Sample size

Target sample size was determined during the planning phase of the project using the mixed effects models simulation routine in PASS 16^102^ which showed that a minimum sample size of 30 per group would provide a power greater than 0.90 to evaluate study hypotheses as the interaction of treatment group with time. Effect sizes for the analysis were based on previous observation of the effects of a similar intervention.^103^

### Procedure

After initial screening, potential participants were schedule for an in-person eligibility visit when, after obtaining verbal consent, they completed measures of health literacy, reading comprehension, a hearing and vision screening, and a medical history interview to determine their eligibility. Eligible persons were then scheduled for a baseline visit during which they completed self-report measures and some individually-administered measures of academic skills. After this baseline visit, participants were randomly assigned to one of the three intervention groups (3^rd^, 6^th^, or 8^th^ grade reading level) and returned to for the intervention visits. These occurred over two to three weeks with a maximum of two sessions per week during which the participants worked through the CDSM modules for a total of three sessions. During intervention sessions, participants worked through the modules on tablet computers (Microsoft Surface Pros®) as preliminary work suggested that many of them would have difficulty in interacting with the modules on smaller screens.

In the first session, participants reviewed an introductory module that explained the purpose of the information, an adherence module that emphasized not only strategies for treatment adherence but also how to work with health care professionals, and a module on stress, its effects, and management techniques. In the second session, participants reviewed modules on sleep, mood, pain, and memory. Finally, in the third session, they worked with modules on fatigue, shortness of breath, and anger.

After completing the modules, participants returned within several days for the first follow-up visit during which they again responded to self-report measures and completed an individual semi-structured interview that elicited their reactions to the modules and the extent to which they had adopted any of the recommendations they contained. Three months later, participants returned for a second follow-up visit during which they again responded to self-report measures and completed the same semi-structured interview.

### Human subjects approval

All study procedures were completed under protocols approved by the Nova Southeastern University Institutional Review Board (2018-685-NSU) and the Emory University Institutional Review Board (MODCR001-IRB00087112). All participants provided verbal consent for screening and written informed consent for all other study procedures.

## Results

Figure 1 presents the CONSORT diagram for participant flow during the trial for both sites combined (separate CONSORT diagrams for each site are provided as supplementary material) and Table 1 presents descriptive data for participants who completed at least one intervention session at each site and for both groups overall. Both participant gender and race were differently distributed at the two sites, with relatively more male and white participants at the Fort Lauderdale site. The two groups of participants did not vary on three of the outcome measures, although Fort Lauderdale participants reported slightly greater medication adherence at a level that approached statistical significance.

**Figure 1.**
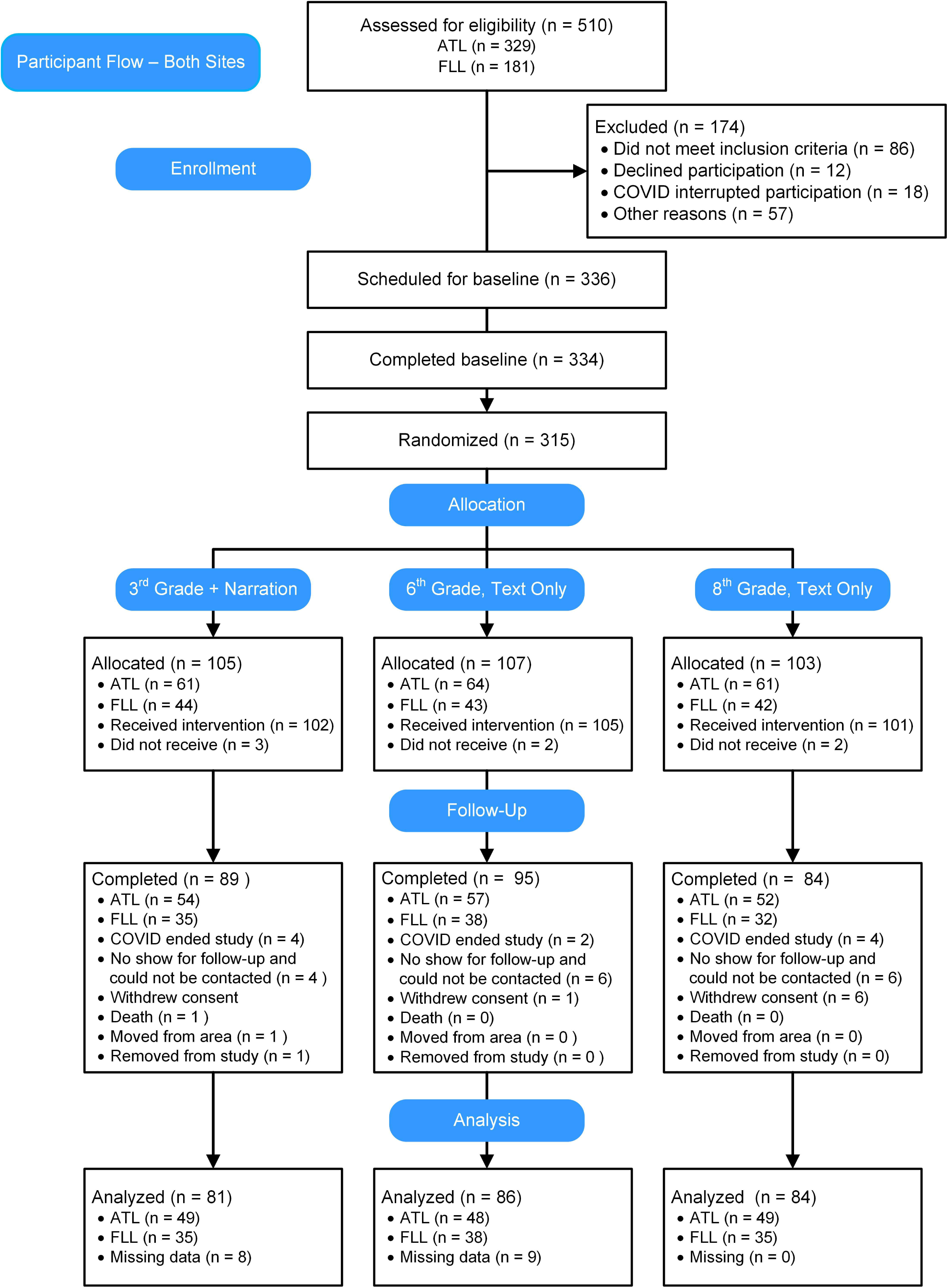
CONSORT diagram.

**Table 1.**
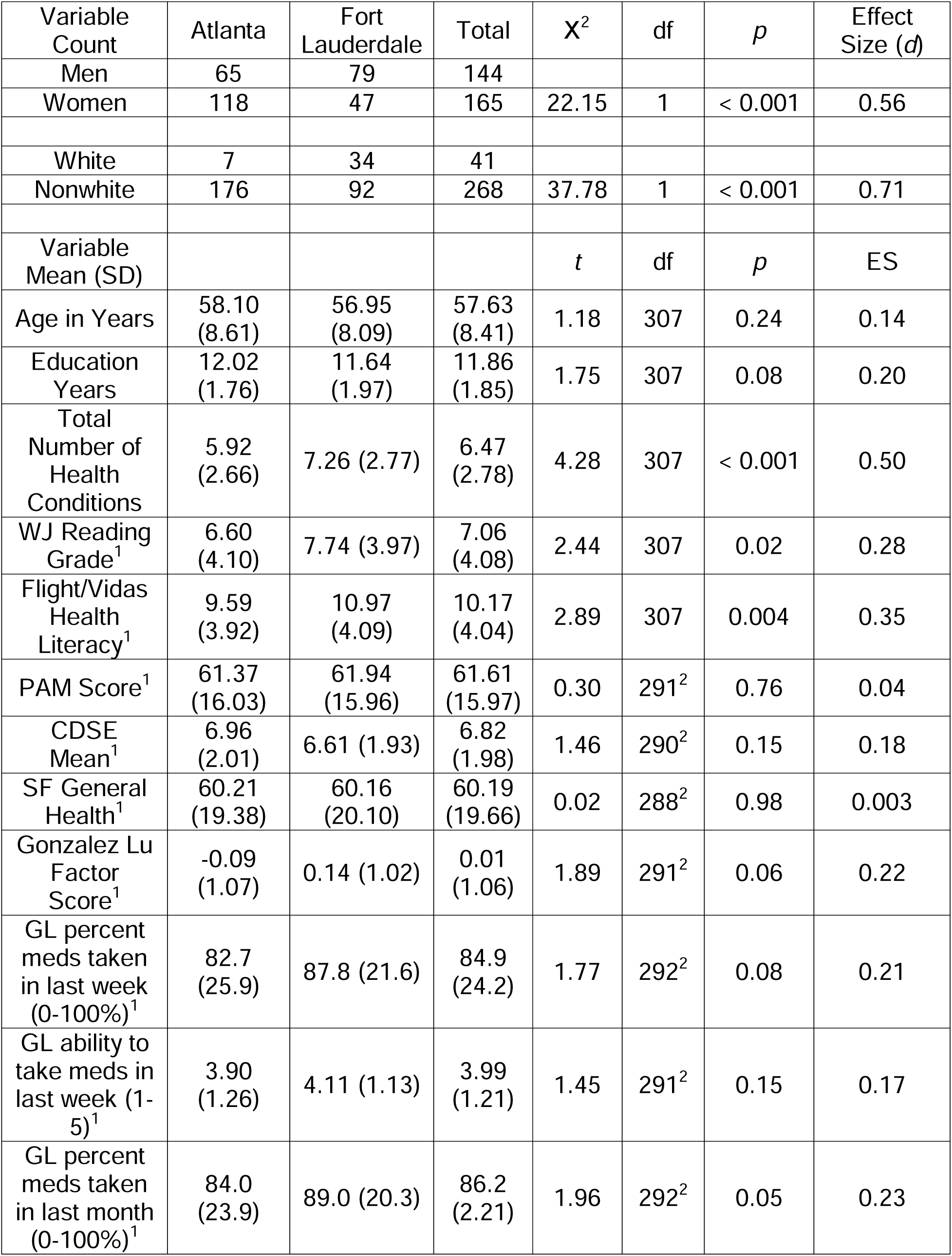
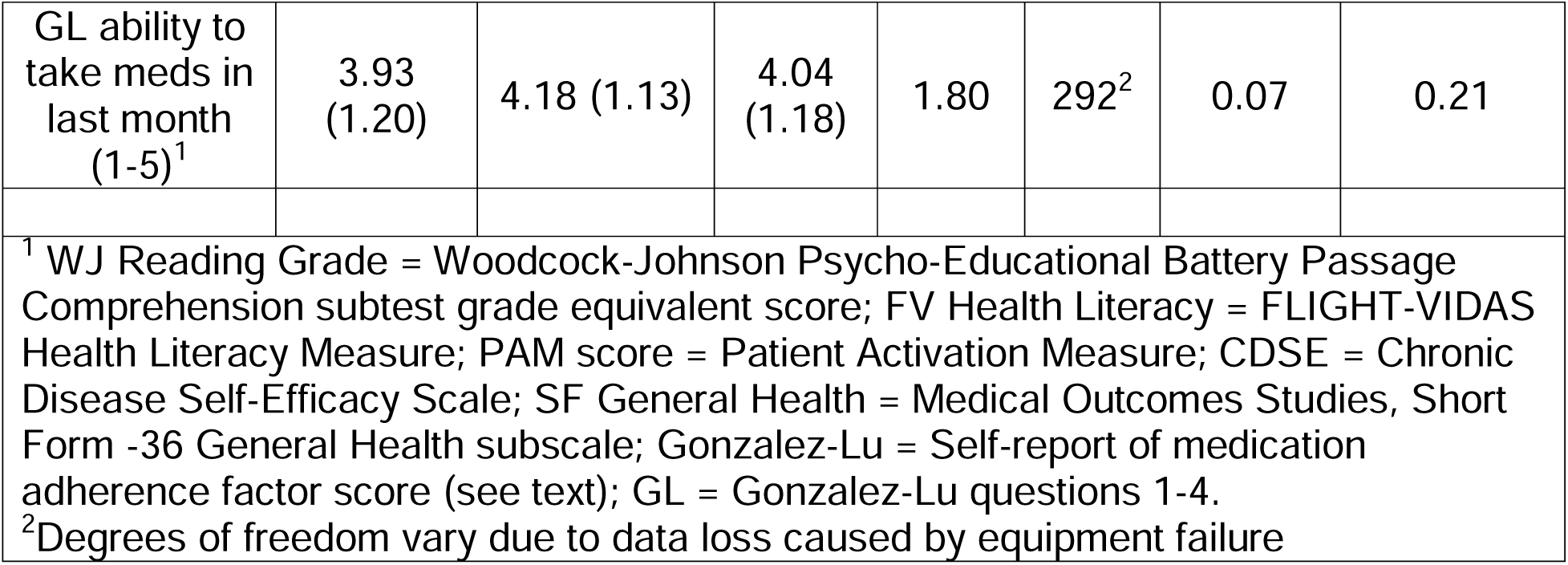
Description of participants completing at least one intervention session.

The random intercept model for the Patient Activation Measure is presented in Table 2, with model-based means for each group at each time displayed in Figure 2. While the interaction of treatment group with time was not statistically significant, there was a significant effect for time, with all groups showing increases in activation after the intervention. Although level of activation appears to continue to increase between the first and second follow-up visits for the 3rd grade group, the difference between this group’s activation and the other groups was not significant (all *p*s > 0.50) and the within-subjects difference also was not significant (*p* > 0.50).

**Table 2.** Model for the Patient Activation Measure.

|  | Sum of<br>Squares | Mean<br>Squares | Numerator<br>df | Denominator<br>df | *F* | *p* |
| --- | --- | --- | --- | --- | --- | --- |
| Age | 601.86 | 601.86 | 1 | 281.16 | 4.90 | **0.03** |
| Gender | 394.9 | 394.9 | 1 | 274.52 | 3.21 | 0.07 |
| Race | 82.26 | 82.26 | 1 | 276.27 | 0.67 | 0.41 |
| Education | 143.46 | 143.46 | 1 | 270.92 | 1.17 | 0.28 |
| Health<br>Literacy | 309.17 | 309.17 | 1 | 276.78 | 2.51 | 0.11 |
| Health<br>Conditions | 189.27 | 189.27 | 1 | 281.6 | 1.54 | 0.22 |
| Site | 17.73 | 17.73 | 1 | 271.62 | 0.14 | 0.70 |
| Time | 822.22 | 411.11 | 2 | 482.77 | 3.34 | **0.04** |
| Group | 159 | 79.5 | 2 | 276.84 | 0.65 | 0.52 |
| Time X<br>Group | 92.61 | 23.15 | 4 | 482.3 | 0.19 | 0.94 |

**Figure 2.**
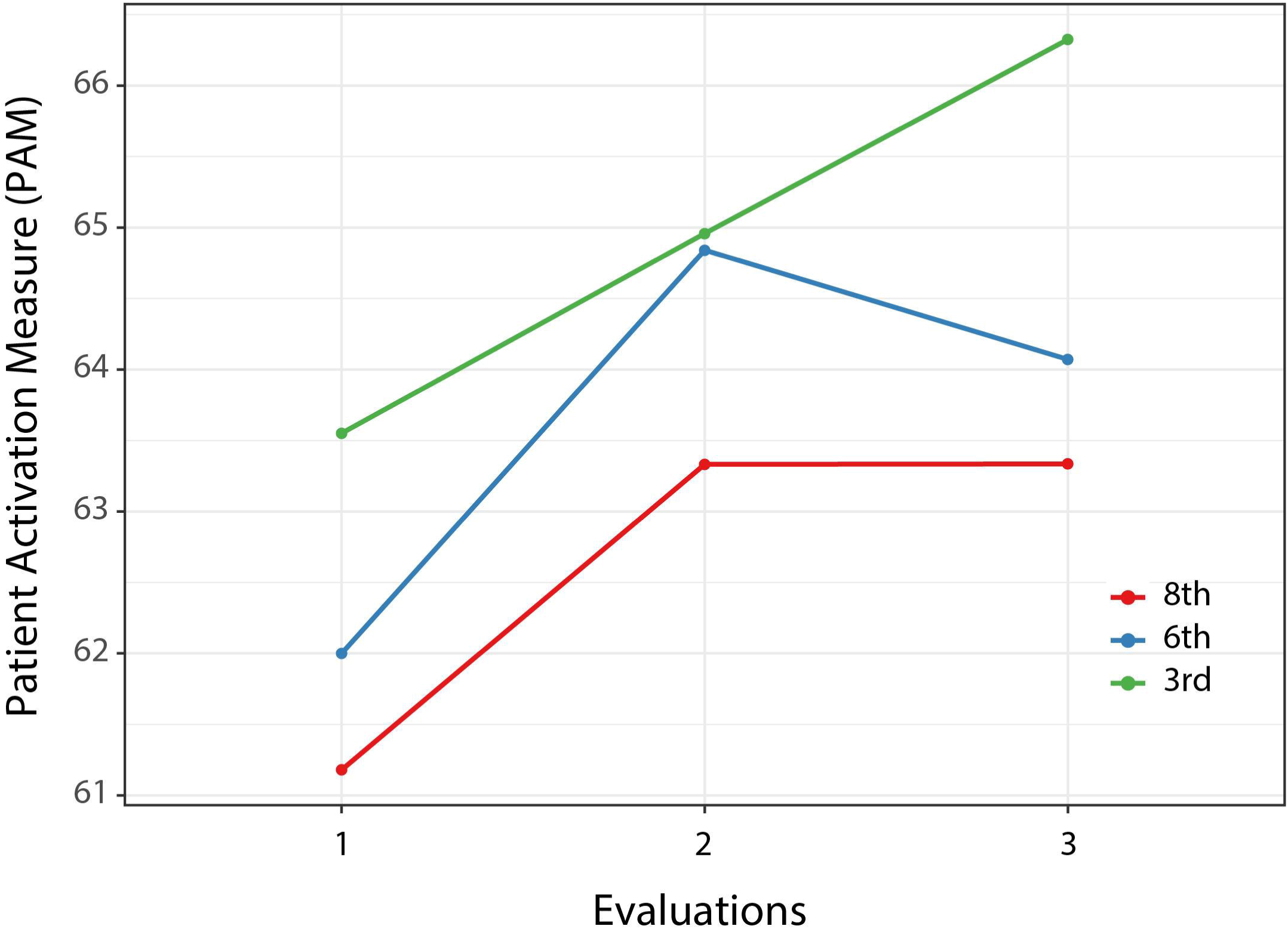
Patient Activation Measure means by group at each evaluation.

The model for the Chronic Disease Self-Efficacy Scale is presented in Table 3. Education, health literacy, and total number of health conditions as well as time were related to this outcome. Model-derived means are plotted in Figure 3. Although the level of self-efficacy appears to decline for the 8^th^ grade group at second follow-up the difference between the 8^th^ and 3^rd^ grade groups (which appears to increase) was not significant (*p* = 0.14).

**Table 3.**
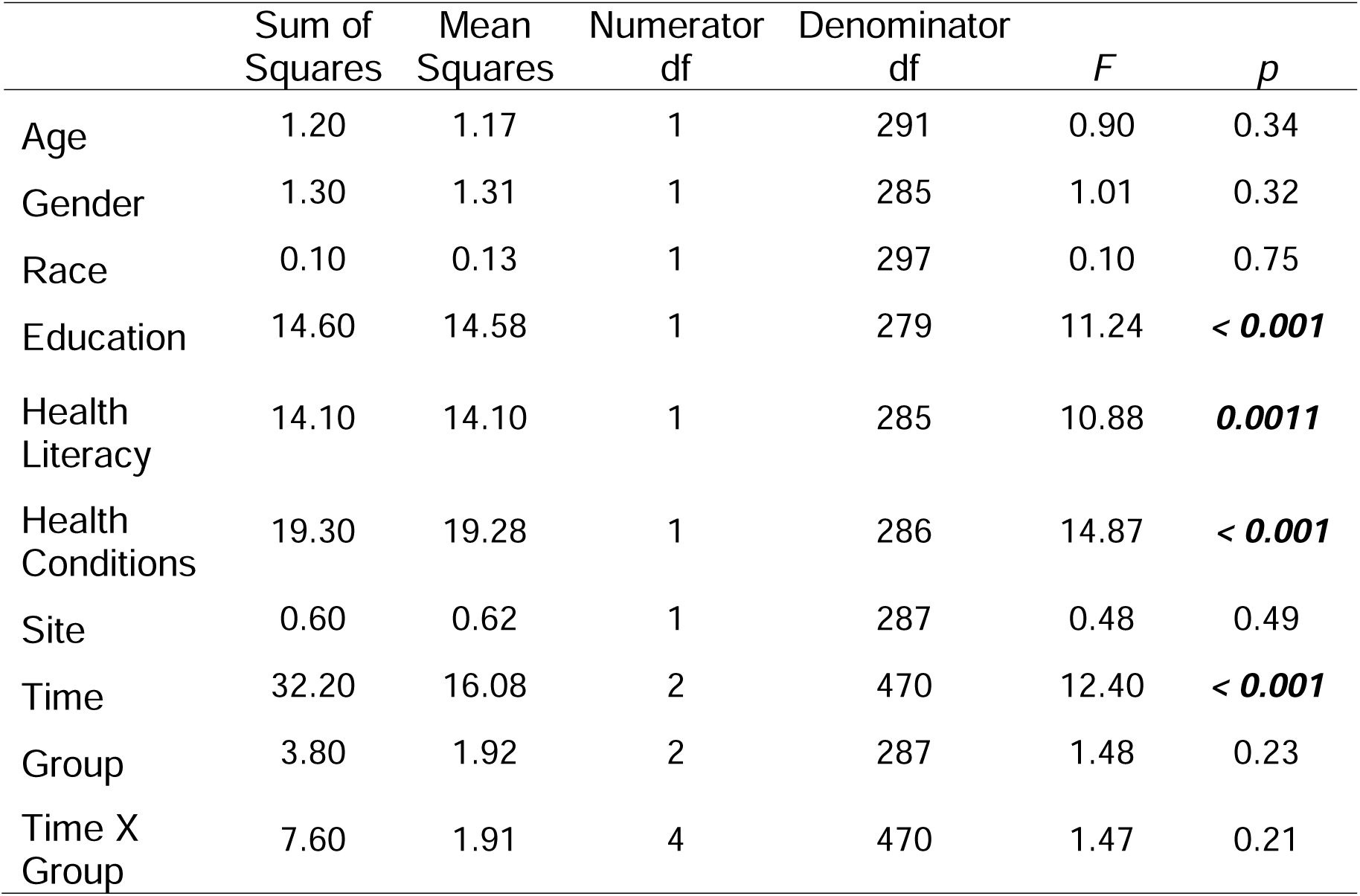
Model for the Chronic Disease Self-Efficacy Scale.

**Figure 3.**
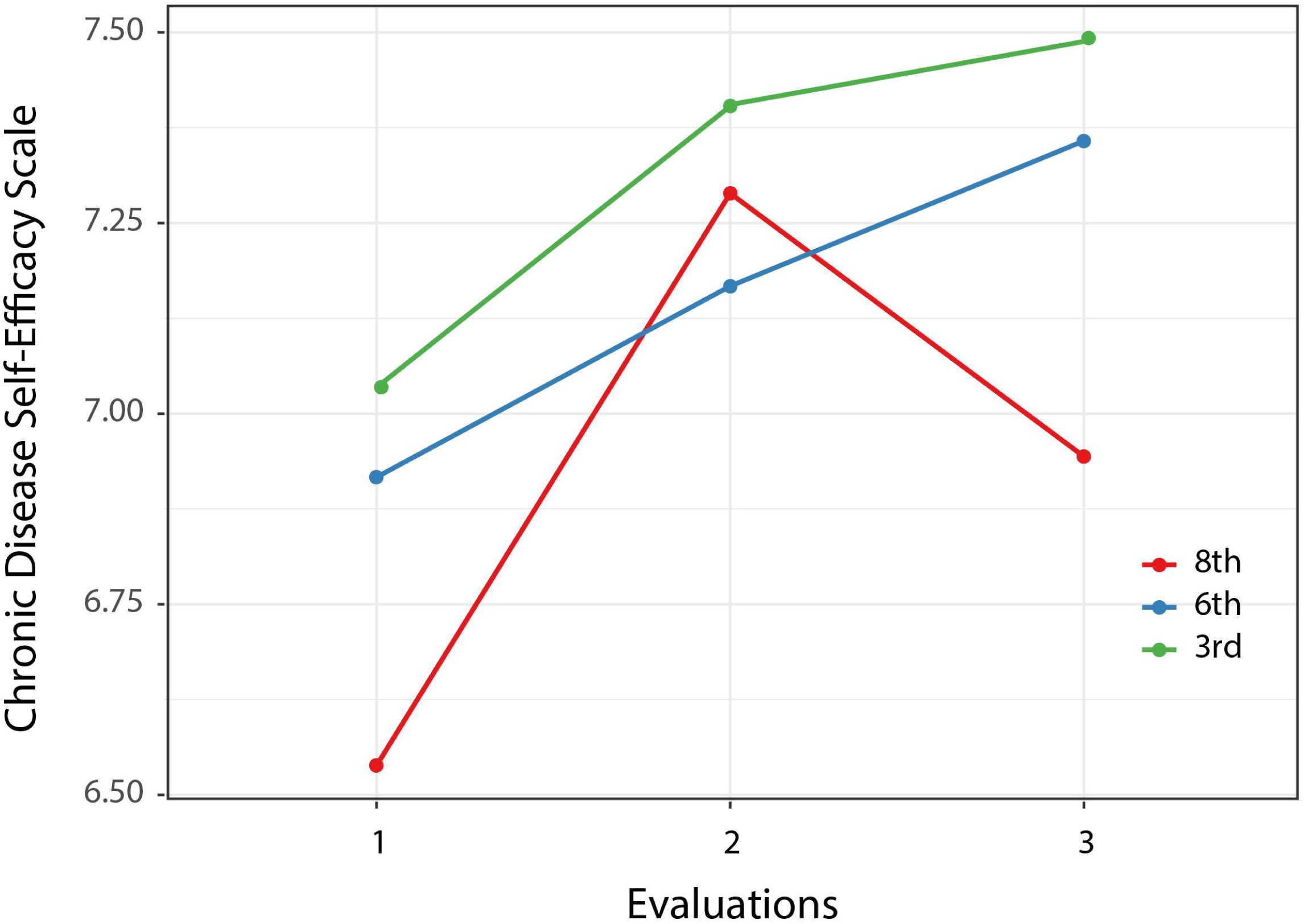
Chronic Disease Self Efficacy means by group at each evaluation.

The model for health-related quality of life (HRQOL; SF-36 General Health scale) is presented in Table 4. In this model, in addition to the effect of time, participants’ HRQOL was positively related to their level of education and inversely related to the number of health conditions they reported. Inspection of Figure 4 shows that the most pronounced effect on this outcome measure was observed at the three-month follow-up. The overall effect for time was associated with a moderate effect size of 0.50. Within group analyses showed that the 8^th^ grade group improved significantly from baseline to the second follow-up (*t* [486] = 2.94, *p* = 0.01), while the 3^rd^ grade group improved significantly only between the first and second follow-up (*t* [486) = 2.66, *p* = 0.02). Although the 6^th^ grade group appears to have improved from the first to second follow-up, this change was not significant (*t* [475] = 1.61, *p* = 0.24).

**Table 4.** Model for MOS SF-36 General Health scale.

|  | Sum of<br>Squares | Mean<br>Squares | Numerator<br>df | Denominator<br>df | *F* | *p* |
| --- | --- | --- | --- | --- | --- | --- |
| Age | 46 | 46 | 1 | 287 | 0.47 | 0.50 |
| Gender | 250 | 250 | 1 | 276 | 2.52 | 0.11 |
| Race | 13 | 13 | 1 | 276 | 0.13 | 0.72 |
| Education | 1139 | 1139 | 1 | 274 | 11.51 | < 0.001 |
| Health<br>Literacy | 259 | 259 | 1 | 277 | 2.62 | 0.11 |
| Health<br>Conditions | 1739 | 1739 | 1 | 280 | 17.57 | < 0.001 |
| Site | 172 | 172 | 1 | 274 | 1.74 | 0.19 |
| Time | 1558 | 779 | 2 | 474 | 7.87 | < 0.001 |
| Group | 272 | 136 | 2 | 277 | 1.37 | 0.26 |
| Time X<br>Group | 355 | 89 | 4 | 474 | 0.9 | 0.47 |

**Figure 4.**
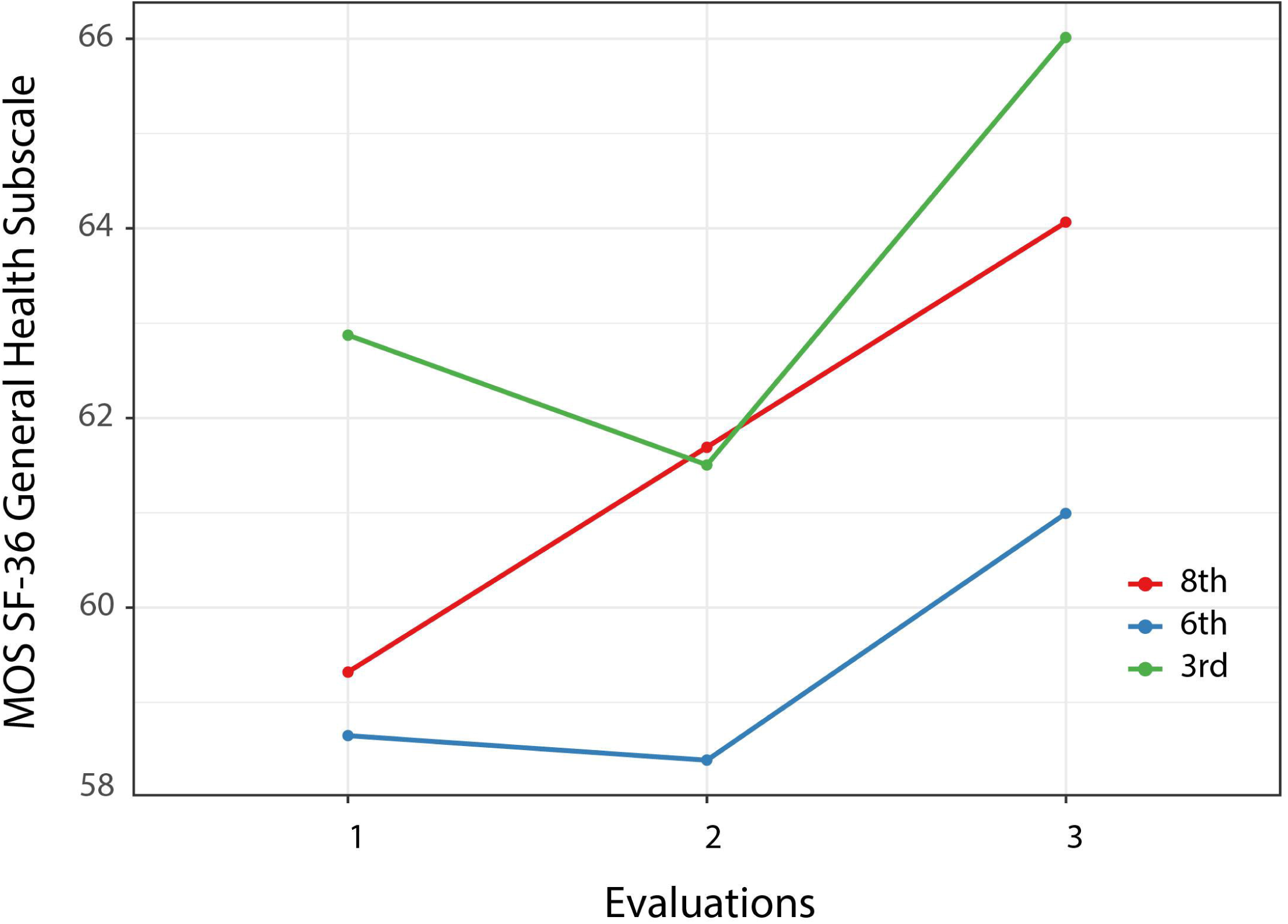
MOS SF-36 General Health means by group at each evaluation.

**Figure 5.**
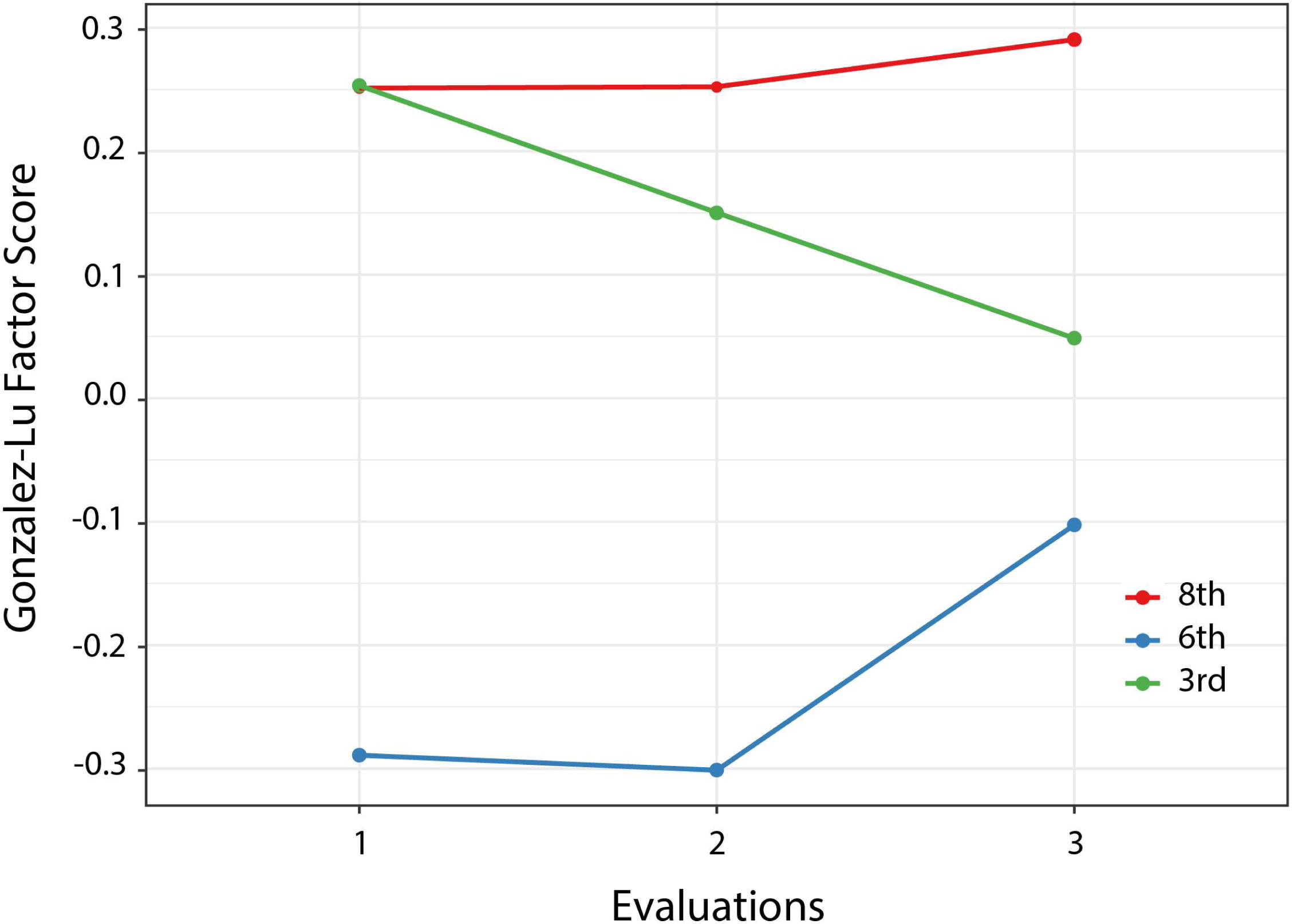
Gonzalez-Lu score means by group at each evaluation.

Finally, the model for self-report medication adherence (Gonzalez-Lu factor score) is presented in Table 5. For medication adherence, no overall effect for time was observed nor was there an interaction of time with treatment group. Both the 8^th^ grade and 3^rd^ grade groups reported significantly greater adherence at baseline (*t* [450] = 3.66, *p* < 0.001) and (*t* [456] = 3.67, *p* < 0.001), respectively. These group differences were maintained at the first follow-up, but only the 8^th^ grade group was still significantly different from the 6^th^ grade group at the second follow-up (*t* [516] = 2.52, *p* = 0.03). Although the 6^th^ grade group appeared to improve adherence between the first and second follow-up, this improvement only approached significance (*t* [477] = 2.07, *p* = 0.10)

**Table 5.** Model for Gonzalez-Lu adherence score.

|  | Sum of<br>Squares | Mean<br>Squares | Numerator<br>df | Denominator<br>df | *F* | *p* |
| --- | --- | --- | --- | --- | --- | --- |
| Age | 1.12 | 1.119 | 1 | 282 | 3.19 | 0.07 |
| Gender | 1.35 | 1.355 | 1 | 274 | 3.87 | 0.0503 |
| Race | 0.63 | 0.633 | 1 | 274 | 1.81 | 0.18 |
| Education | 0.01 | 0.008 | 1 | 270 | 0.02 | 0.88 |
| Health<br>Literacy | 1.61 | 1.609 | 1 | 275 | 4.59 | 0.03 |
| Health<br>Conditions | 0.94 | 0.94 | 1 | 280 | 2.68 | 0.10 |
| Site | 0.79 | 0.786 | 1 | 272 | 2.24 | 0.14 |
| Time | 0.28 | 0.138 | 2 | 476 | 0.39 | 0.67 |
| Group | 5.73 | 2.864 | 2 | 276 | 8.18 | <0.001 |
| Time X<br>Group | 3.09 | 0.772 | 4 | 476 | 2.2 | 0.07 |

## Discussion

The purpose of this study was to investigate the impact of a tailored information app for CDSM in older persons chronic health conditions and low health literacy. We hypothesized that the app would have a positive effect on four outcomes that reflect patients’ attitudes and beliefs about their ability to manage their health, activation, self-efficacy, HRQOL, and medication adherence. Results partially support this hypothesis, as we saw a significant positive impact of the app on three of the outcomes, activation, self-efficacy, and health-related quality of life. We did not find a significant change in participants’ self-report of medication adherence.

Outcome measures for this study were chosen because of their relevance to older person’s ability to manage their health and their use in previous studies, allowing us to compare results of the automated app with those of similar in-person and internet-delivered CDSM interventions. Other researchers, for example, have found that CDSM programs can have a positive impact on self-efficacy for disease management.^41, 45, 47^ One trial of delivering a CDSM intervention via the internet^46^ did not include self-efficacy as an outcome. A number of studies, however, have included self-reported general health (here we have termed it health-related quality of life or HRQOL) as an outcome, frequently finding positive effects of a CDSM intervention including a trial of delivering it via the internet.^46^ Improved health-related quality of life was observed in this study as well, associated with a moderate effect size (0.50) that has been described as clinically meaningful for QOL outcomes in other areas.^104^

These results are thus similar to those observed in studies of in-person CDSM programs that have been shown to have a positive impact on activation,^105^ self-efficacy, and HRQOL. This suggests that some of the drawbacks to providing in-person CDSM interventions (cost, lack of trained personnel, accessibility) may be addressed by providing patients access to CDSM as a digital therapeutic. While it is true that the development of the mobile app was expensive initially, after initial deployment this sort of app can be inexpensive to maintain while providing access to large numbers of patients. We^106^ and others^107^ have shown that initial development and ongoing deployment costs can be substantially offset by benefits such as improved self-management behavior. Lindsay et al.,^61^ for example, showed that a substantial increase in activation was associated with lower follow-up costs, especially in high risk populations.

Given the common critique of applications that do not provide patients information at the 3^rd^ to 6^th^ grade level, it is not clear why we did not find an effect of the modules’ text difficulty favoring the two experimental groups (3^rd^ and 6^th^ grade text) over the control (8^th^ grade text). A possible explanation comes from research on educational applications in other situations in which researchers found a “contiguity effect”^108^ in which learners presented text and graphics in close proximity, as was done in this study, resulted in superior learning compared to a condition in which graphic and text elements were separated. In more recent studies, research has shown that use of multimedia as an adjunct to traditional instruction may enhance comprehension and learner motivation.^109^ Other research has shown that incorporating information technology in interventions may enhance patient engagement,^110^ an effect that was not controlled in our research design. Future research may help clarify whether multimedia instruction can help persons with low health literacy even when the text of the information presented is too difficult.

Limitations of this study include the nature of the sample, which was nonwhite in the majority, and differences between the two study sites on some variables. While we included variables on which the sites differed as covariates in statistical models as well as using site as an additional covariate, it is possible that the observed differences may have affected the study’s outcomes. On the other hand, in our search for persons with low health literacy, we succeeded in recruiting a large number of participants health literacy skills that might place them at a disadvantage in managing their health, and the study shows that the intervention was successful in increasing activation, self-efficacy, and HRQOL. The lack of between-site differences on outcome variables also supports the usefulness of the intervention, as it appears to have been efficacious in two diverse settings. A significant limitation is the lack of a finding on control vs experimental group outcomes. Although lack of differences related to reading difficulty is a concern, the clear effect of time suggests that the intervention had a significant impact on participants. Even without significant between-group differences, the alternative that participants improved in activation, self-efficacy, and HRQOL spontaneously over time is implausible. Additional research on possible mediators and moderators of change in these measures would be useful in further understanding these finding and enhancing the efficacy of the app.

## Conclusions

In this study we investigated the effects of a mobile app for CDSM in older persons with low levels of health literacy and chronic health conditions. Although we hypothesized that modules that presented content at the 8^th^ grade level would be less effective than those at lower levels, this hypothesis was not supported. Clear effects of the intervention over time, however, suggest that it had positive effects for all groups. Future development of the app will include additional analyses of possible mediators and moderators of its effects to better understand how it works and ultimately have an even greater impact.

## Data Availability

All data produced in the present study are available upon reasonable request to the authors.

## Ethics approval

This study was approved by the Institutional Review Boards of Emory University (MODCR001-IRB00087112) and of Nova Southeastern University (2018-685-NSU)

## Notes

Roles of authors: RLO analyzed the data and drafted the manuscript. RLO, DW, MS, RD, JC, KK, and KTP contributed to conceptualization, investigation, methods, validation; RLO and DW were responsible for funding acquisition; RLO, RD, VA, JS, NP and DP conducted data curation, supervision, and project administration. All authors read and approved the final manuscript.

Guarantor: RLO

## References

1. US Department of Health and Human Services. Healthy people 2030: Objectives and data. Washington DC: Department of Health and Human Services. Available at https://health.gov/healthypeople/objectives-and-dataAccessed March 1, 2023, 2021.

2. Kutner M, Greenberg E, Jin Y, et al. The health literacy of America’s adults: Results from the 2003 National Asessment of Adult Literacy (NCES 2006-483). Washington, DC: US Department of Education, National Center for Educational Statistics, 2006.

3. OECD. OECD skills outlook 2013: First results from the survey of adults skills. Paris: OECD Publishing, 2013.

4. National Center for Educational Statistics. US adults with low literacy and numeracy skills: 2012/2014 to 2017. Washington, DC: National Center for Educational Statistics, 2022.

5. Baccolini V, Rosso A, Di Paolo C, et al. What is the prevalence of low health literacy in European Union member states? A systematic review and meta-analysis. J Gen Intern Med 2021; 36: 753–761. 20210105. DOI: 10.1007/s11606-020-06407-8.

6. Canadian Council on Learning. Health literacy in Canada: A healthy understanding. Ottawa: Canadian Council on Learning, 2008.

7. Adekoya-Cole TO, Akinmokun OI, Enweluzo GO, et al. Poor health literacy in Nigeria: Causes, consequences and measures to improve it. Nig Q J Hosp Med 2015; 25: 112–117.

8. Paasche-Orlow MK and Wolf MS. Promoting health literacy research to reduce health disparities. J Health Commun 2010; 15 Suppl 2: 34–41. DOI: 926961883 [pii];10.1080/10810730.2010.499994 [doi].

9. Stormacq C, Van den Broucke S and Wosinski J. Does health literacy mediate the relationship between socioeconomic status and health disparities? Integrative review. Health Promot Int 2019; 34: e1–e17. 2018/08/15. DOI: 10.1093/heapro/day062.

10. Mantwill S, Monestel-Umana S and Schulz PJ. The relationship between health literacy and health disparities: A systematic review. PLoS One 2015; 10: e0145455. 2015/12/25. DOI: 10.1371/journal.pone.0145455.

11. Pignone M, Dewalt DA, Sheridan S, et al. Interventions to improve health outcomes for patients with low literacy. A systematic review. J Gen Intern Med 2005; 20: 185–192. DOI: JGI40208 [pii];10.1111/j.1525-1497.2005.40208.x [doi].

12. Berkman ND, Sheridan SL, Donahue KE, et al. Low health literacy and health outcomes: an updated systematic review. Ann Intern Med 2011; 155: 97–107. DOI: 155/2/97 [pii];10.1059/0003-4819-155-2-201107190-00005 [doi].

13. Bo A, Friis K, Osborne RH, et al. National indicators of health literacy: ability to understand health information and to engage actively with healthcare providers - a population-based survey among Danish adults. BMC Public Health 2014; 14: 1095. 2014/10/24. DOI: 10.1186/1471-2458-14-1095.

14. Osborn CY, Cavanaugh K, Wallston KA, et al. Health literacy explains racial disparities in diabetes medication adherence. J Health Commun 2011; 16 Suppl 3: 268–278. DOI: 10.1080/10810730.2011.604388 [doi].

15. Osborn CY, Cavanaugh K, Wallston KA, et al. Diabetes numeracy: an overlooked factor in understanding racial disparities in glycemic control. Diabetes Care 2009; 32: 1614–1619. DOI: dc09-0425 [pii];10.2337/dc09-0425 [doi].

16. Osborn CY, Paasche-Orlow MK, Bailey SC, et al. The mechanisms linking health literacy to behavior and health status. Am J Health Behav 2011; 35: 118–128. DOI: 10.5555/ajhb.2011.35.1.118 [pii].

17. Osborn CY, Paasche-Orlow MK, Davis TC, et al. Health literacy: An overlooked factor in understanding HIV health disparities. Am J Prev Med 2007; 33: 374–378. DOI: S0749-3797(07)00465-5 [pii];10.1016/j.amepre.2007.07.022 [doi].

18. Waldrop-Valverde D, Osborn CY, Rodriguez A, et al. Numeracy skills explain racial differences in HIV medication management. AIDS Behav 2010; 14: 799–806. DOI: 10.1007/s10461-009-9604-4 [doi].

19. Rowlands G, Protheroe J, Winkley J, et al. A mismatch between population health literacy and the complexity of health information: An observational study. Br J Gen Pract 2015; 65: e379–386. DOI: 10.3399/bjgp15X685285.

20. Kessels RPC. Patients’ memory for medical information. Journal of the Royal Society of Medicine 2003; 96: 219–222.

21. Kravitz RL, Hays RD, Sherbourne CD, et al. Recall of recommendations and adherence to advice among patients with chronic medical conditions. Arch Intern Med 1993; 153: 1869–1878.

22. Vinker S, Eliyahu V and Yaphe J. The effect of drug information leaflets on patient behavior. Isr Med Assoc J 2007; 9: 383–386.

23. Herber OR, Gies V, Schwappach D, et al. Patient information leaflets: Informing or frightening? A focus group study exploring patients’ emotional reactions and subsequent behavior towards package leaflets of commonly prescribed medications in family practices. BMC Family Practice 2014; 15: 163. DOI: 10.1186/1471-2296-15-163.

24. Kreuter M, Farrell D, Olevitch L, et al. Tailoring health messages. Mahwah, NJ: Lawrence Erlbaum, 2000.

25. Schapira MM, Swartz S, Ganschow PS, et al. Tailoring educational and behavioral interventions to level of health literacy: A systematic review. MDM Policy Pract 2017; 2: 2381468317714474. 20170615. DOI: 10.1177/2381468317714474.

26. Taggart J, Williams A, Dennis S, et al. A systematic review of interventions in primary care to improve health literacy for chronic disease behavioral risk factors. BMC Family Practice 2012; 13: 49. DOI: 10.1186/1471-2296-13-49.

27. Berkman ND, Sheridan SL, Donahue KE, et al. Health literacy interventions and outcomes: an updated systematic review. Evidence report/technology assesment no. 199. Rockville, MD: Agency for Healthcare Research and Quality, 2011.

28. Schillinger D, Duran ND, McNamara DS, et al. Precision communication: Physicians’ linguistic adaptation to patients’ health literacy. Science Advances 2021; 7: eabj2836. DOI: doi:10.1126/sciadv.abj2836.

29. Ghalibaf AK, Nazari E, Gholian-Aval M, et al. Comprehensive overview of computer-based health information tailoring: A systematic scoping review. BMJ Open 2019; 9: e021022. DOI: 10.1136/bmjopen-2017-021022.

30. Lustria ML, Noar SM, Cortese J, et al. A meta-analysis of web-delivered tailored health behavior change interventions. J Health Commun 2013; 18: 1039–1069. DOI: 10.1080/10810730.2013.768727 [doi].

31. Ownby RL, Acevedo A and Waldrop-Valverde D. Enhancing the impact of mobile health literacy interventions to reduce health disparities. Quarterly Review of Distance Education 2019; 10: 15–34.

32. Ginsburg GS and Phillips KA. Precision medicine: From science to value. Health Aff (Millwood) 2018; 37: 694–701. DOI: 10.1377/hlthaff.2017.1624.

33. Centers for Disease Control. CDC clear communication index user guide. Atlanta GA: Centers for Disease Control, 2019.

34. US Department of Health and Human Services. Healthy people 2020: Topics and objectives. Washington DC: Department of Health and Human Services. Available at http://healthypeople.gov/2020/topicsobjectives2020/pdfs/HP2020objectives.pdf. Accessed August 1, 2011, 2011.

35. Protheroe J and Rowlands G. Matching clinical information with levels of patient health literacy. Nurs Manag (Harrow) 2013; 20: 20–21. DOI: 10.7748/nm2013.06.20.3.20.e1095.

36. Eltorai AE, Ghanian S, Adams CA, Jr., et al. Readability of patient education materials on the American Association for Surgery of Trauma website. Arch Trauma Res 2014; 3: e18161. 20140430. DOI: 10.5812/atr.18161.

37. Hutchinson N, Baird GL and Garg M. Examining the reading level of internet medical information for common internal medicine diagnoses. The American Journal of Medicine 2016; 129: 637–639. DOI: 10.1016/j.amjmed.2016.01.008.

38. Miles RC, Baird GL, Choi P, et al. Readability of online patient educational materials related to breast lesions requiring surgery. Radiology 2019; 291: 112–118. DOI: 10.1148/radiol.2019182082.

39. Rooney MK, Santiago G, Perni S, et al. Readability of patient education materials From high-impact medical journals: A 20-year analysis. J Patient Exp 2021; 8: 2374373521998847. 20210303. DOI: 10.1177/2374373521998847.

40. Ownby RL. Readability of consumer-oriented geriatric depression information on the internet. Clinical Gerontologist 2006; 29: 17–32. DOI: 10.1300/J018v29n04_02.

41. Lorig KR, Sobel DS, Ritter PL, et al. Effect of a self-management program on patients with chronic disease. Eff Clin Pract 2001; 4: 256–262.

42. Ownby RL, Acevedo A, Waldrop-Valverde D, et al. A mobile app for chronic disease self-management: Protocol for a randomized controlled trial. JMIR Res Protoc 2017; 6: e53. DOI: 10.2196/resprot.7272.

43. Bodenheimer T, Lorig K, Holman H, et al. Patient self-management of chronic disease in primary care. JAMA 2002; 288: 2469–2475. 2002/11/19.

44. Lorig K, Ritter PL, Ory MG, et al. Effectiveness of a generic chronic disease self-management program for people with type 2 diabetes: A translation study. Diabetes Educ 2013. DOI: 0145721713492567 [pii];10.1177/0145721713492567 [doi].

45. Lorig KR, Ritter P, Stewart AL, et al. Chronic disease self-management program: 2-year health status and health care utilization outcomes. Med Care 2001; 39: 1217–1223.

46. Lorig KR, Ritter PL, Laurent DD, et al. Internet-based chronic disease self-management: A randomized trial. Med Care 2006; 44: 964–971. DOI: 10.1097/01.mlr.0000233678.80203.c1 [doi];00005650-200611000-00002 [pii].

47. Ory MG, Ahn S, Jiang L, et al. Successes of a national study of the Chronic Disease Self-Management Program: Meeting the triple aim of health care reform. Med Care 2013; 51: 992–998. DOI: 10.1097/MLR.0b013e3182a95dd1.

48. Jaglal S, Guilcher S, Hawker G, et al. Impact of a chronic disease self-management program on health care utilization in rural communities: a retrospective cohort study using linked administrative data. BMC Health Services Research 2014; 14: 198. DOI: 10.1186/1472-6963-14-198.

49. Stormacq C, Wosinski J, Boillat E, et al. Effects of health literacy interventions on health-related outcomes in socioeconomically disadvantaged adults living in the community: A systematic review. JBI Evid Synth 2020; 18: 1389–1469. DOI: 10.11124/jbisrir-d-18-00023.

50. Lewis D. Computer-based approaches to patient education: a review of the literature. J Am Med Inform Assoc 1999; 6: 272–282.

51. Lustria ML, Cortese J, Noar SM, et al. Computer-tailored health interventions delivered over the Web: Review and analysis of key components. Patient Educ Couns 2009; 74: 156–173. DOI: S0738-3991(08)00469-2 [pii];10.1016/j.pec.2008.08.023 [doi].

52. Kuhn E, Weiss BJ, Taylor KL, et al. CBT-I Coach: A description and clinician perceptions of a mobile app for cognitive behavioral therapy for insomnia. Journal of clinical sleep medicine 2016; 12: 597–606. DOI: 10.5664/jcsm.5700.

53. McNaughton JL. Brief interventions for depression in primary care: a systematic review. Can Fam Physician 2009; 55: 789–796. DOI: 55/8/789 [pii].

54. Hibbard JH, Stockard J, Mahoney ER, et al. Development of the Patient Activation Measure (PAM): conceptualizing and measuring activation in patients and consumers. Health Serv Res 2004; 39: 1005–1026. DOI: 10.1111/j.1475-6773.2004.00269.x [doi];HESR269 [pii].

55. Greene J and Hibbard JH. Why does patient activation matter? An examination of the relationships between patient activation and health-related outcomes. J Gen Intern Med 2012; 27: 520–526. DOI: 10.1007/s11606-011-1931-2.

56. Hosseinzadeh H, Verma I and Gopaldasani V. Patient activation and Type 2 diabetes mellitus self-management: A systematic review and meta-analysis. Australian Journal of Primary Health 2020; 26: 431–442. DOI: https://doi.org/10.1071/PY19204.

57. Mosen DM, Schmittdiel J, Hibbard J, et al. Is patient activation associated with outcomes of care for adults with chronic conditions? The Journal of Ambulatory Care Management 2007; 30: 21–29.

58. Hibbard J and Greene J. What the evidence shows about patient activation: Better health outcomes and care experiences; fewer eata on costs. Health Affairs 2013; 32: 207–214. DOI: 10.1377/hlthaff.2012.1061.

59. Greene J, Hibbard J and Tusler M. How much do health literacy and patient activation contribute to older adults’ ability to manage their health? Washington, DC: AARP. Available at: http://assets.aarp.org/rgcenter/health/2005_05_literacy.pdf, 2005.

60. Greene J, Hibbard JH, Sacks R, et al. When patient activation levels change, health outcomes and costs change, too. Health Aff (Millwood) 2015; 34: 431–437. DOI: 10.1377/hlthaff.2014.0452.

61. Lindsay A, Hibbard JH, Boothroyd DB, et al. Patient activation changes as a potential signal for changes in health care costs: Cohort study of US high-cost patients. Journal of General Internal Medicine 2018; 33: 2106–2112. DOI: 10.1007/s11606-018-4657-6.

62. Bandura A. The primacy of self-regulation in health promotion. Applied Psychology 2005; 54: 245–254. DOI: https://doi.org/10.1111/j.1464-0597.2005.00208.x.

63. Bandura A. Health promotion from the perspective of social cognitive theory. Psychology & Health. Routledge, 1998, p. 623–649.

64. Chen J, Tian Y, Yin M, et al. Relationship between self-efficacy and adherence to self-management and medication among patients with chronic diseases in China: A multicentre cross-sectional study. Journal of Psychosomatic Research 2023; 164: 111105. DOI: https://doi.org/10.1016/j.jpsychores.2022.111105.

65. Hoong JM, Koh HA, Wong K, et al. Effects of a community-based chronic disease self-management programme on chronic disease patients in Singapore. Chronic Illness; 0: 17423953221089307. DOI: 10.1177/17423953221089307.

66. Farley H. Promoting self-efficacy in patients with chronic disease beyond traditional education: A literature review. Nurs Open 2020; 7: 30–41. 20191020. DOI: 10.1002/nop2.382.

67. Fernandez-Lazaro CI, García-González JM, Adams DP, et al. Adherence to treatment and related factors among patients with chronic conditions in primary care: A cross-sectional study. BMC Family Practice 2019; 20: 132. DOI: 10.1186/s12875-019-1019-3.

68. DiMatteo MR. Variations in patients’ adherence to medical recommendations: A quantitative review of 50 years of research. Med Care 2004; 42: 200–209. 2004/04/13. DOI: 10.1097/01.mlr.0000114908.90348.f9.

69. Walsh CA, Cahir C, Tecklenborg S, et al. The association between medication non-adherence and adverse health outcomes in ageing populations: A systematic review and meta-analysis. British Journal of Clinical Pharmacology 2019; 85: 2464–2478. DOI: https://doi.org/10.1111/bcp.14075.

70. Cutler RL, Fernandez-Llimos F, Frommer M, et al. Economic impact of medication non-adherence by disease groups: A systematic review. BMJ Open 2018; 8: e016982. 20180121. DOI: 10.1136/bmjopen-2017-016982.

71. Jacobs RJ, Ownby RL, Acevedo A, et al. A qualitative study examining health literacy and chronic illness self-management in Hispanic and non-Hispanic older adults. J Multidiscip Healthc 2017; 10: 167–177. 2017/05/04. DOI: 10.2147/JMDH.S135370.

72. Paas F, Renkl A and Sweller J. Cognitive Load Theory: Instructional implications of the interaction between information structures and cognitive architecture. Instructional Science 2004; 32: 1–8.

73. Van Merrienboer JJ and Sweller J. Cognitive load theory in health professional education: Design principles and strategies. Med Educ 2010; 44: 85–93. DOI: MED3498 [pii];10.1111/j.1365-2923.2009.03498.x [doi].

74. Mayer RE. Multimedia learning (2nd ed.). New York: Cambridge, 2009.

75. Patel N, Waldrop D and Ownby RL. Creating a tailored info app to promote self-management skills in persons with chronic health conditions: Development strategies and user experience [preprint] PsyArXiv 2023, January 26. DOI: 10.31234/osf.io/98bxk.

76. Fry E. A readability formula that saves time. Journal of Reading 1968; 11: 513–578.

77. Flesch R. A new readability yardstick. Journal of Applied Psychology 1948; 32: 221–233.

78. Kincaid JP, Fishburne RP, Rogers RL, et al. Derivation of new readability formulas (Automated Readability Index, Fog Count, and Flesch Reading Ease Formula) for Navy enlisted personnel. Millington, TN: Chief of Naval Technical Training. Available online at http://www.dtic.mil/dtic/tr/fulltext/u2/a006655.pdf., 1975.

79. National Institutes of Health Staff. What are the requirements for reporting on human study participant age, sex/gender, and race/ethnicity?, https://nexus.od.nih.gov/all/2021/04/29/what-are-the-requirements-for-reporting-on-human-study-participant-age-sex-gender-and-race-ethnicity/ (2021, accessed March 28, 2023 2023).

80. Murphy PW, Davis TC, Long SW, et al. Rapid Estimate of Adult Literacy in Medicine (REALM): A quick reading test for patients. Journal of Reading 1993; 37: 124–130.

81. Han HJ, Acevedo A, Waldrop-Valverde D, et al. Evaluation of short forms of the Rapid Estimate of Adult Literacy in Medicine (REALM). International Conference on Communication in Healthcare/Health Literacy Annual Researach Conference. Baltimore MD2017, October.

82. Groll DL, To T, Bombardier C, et al. The development of a comorbidity index with physical function as the outcome. Journal of clinical epidemiology 2005; 58: 595–602.

83. Centers for Medicare and Medicaid Services. Chronic conditions among Medicare beneficiaries, chartbook (2012 ed.). Baltimore, MD: Centers for Medicare and Medicaid Services, 2012.

84. Woodcock RW, McGrew KS and Mather N. Woodcock-Johnson III Normative Update. Rolling Meadows, IL: Riverside, 2007.

85. Ownby RL, Acevedo A, Waldrop-Valverde D, et al. Development and initial validation of a computer-administered health literacy assessment in Spanish and English: FLIGHT/VIDAS. Patient Related Outcome Measures 2013; 4: 1–15.

86. Pleasant A. Advancing health literacy measurement: a pathway to better health and health system performance. J Health Commun 2014; 19: 1481–1496. DOI: 10.1080/10810730.2014.954083 [doi].

87. Hibbard JH, Mahoney ER, Stockard J, et al. Development and testing of a short form of the Patient Activation Measure. Health Serv Res 2005; 40: 1918–1930. DOI: 10.1111/j.1475-6773.2005.00438.x.

88. Skolasky RL, Green AF, Scharfstein D, et al. Psychometric properties of the patient activation measure among multimorbid older adults. Health Serv Res 2011; 46: 457–478. 20101119. DOI: 10.1111/j.1475-6773.2010.01210.x.

89. Lorig K, Stewart A, Ritter P, et al. Outcome measures for health education and other health care interventions. Thousand Oaks CA: Sage, 1996.

90. Ware JE and Sherbourne CD. The MOS 36-item short-form health survey (SF-36). I. Conceptual framework and item selection. Med Care 1992; 30: 478–483.

91. Peeters G, Waller M and Dobson AJ. SF-36 normative values according to level of functioning in older women. Quality of Life Research 2019; 28: 979–989. DOI: https://doi.org/10.1007/s11136-018-2077-z.

92. Lyons RA, Perry IM and Littlepage BNC. Evidence for the validity of the Short-form 36 Questionnaire (SF-36) in an elderly population. Age and Ageing 1994; 23: 182–184. DOI: 10.1093/ageing/23.3.182.

93. Butterly EW, Hanlon P, Shah ASV, et al. Comorbidity and health-related quality of life in people with a chronic medical condition in randomised clinical trials: An individual participant data meta-analysis. PLOS Medicine 2023; 20: e1004154. DOI: 10.1371/journal.pmed.1004154.

94. Gonzalez JS, Schneider HE, Wexler DJ, et al. Validity of medication adherence self-reports in adults with type 2 diabetes. Diabetes Care 2013; 36: 831–837. DOI: dc12-0410 [pii];10.2337/dc12-0410 [doi].

95. Lu M, Safren SA, Skolnik PR, et al. Optimal recall period and response task for self-reported HIV medication adherence. AIDS Behav 2008; 12: 86–94. DOI: 10.1007/s10461-007-9261-4 [doi].

96. McNeish D and Wolf MG. Thinking twice about sum scores. Behavior Research Methods 2020; 52: 2287–2305. DOI: 10.3758/s13428-020-01398-0.

97. Ferrando PJ and Lorenzo-Seva U. On the added value of multiple factor score estimates in essentially unidimensional models. Educ Psychol Meas 2019; 79: 249–271. 20180515. DOI: 10.1177/0013164418773851.

98. R Core Team. R: A language and environment for statistical computing. Vienna, Austria: R foundation for statistical computing, 2022.

99. Bates D, Maechler M, Bolker B, et al. Fitting linear mixed-effects models using lme4. Journal of Statistical Software 2015; 67: 1–48. DOI: doi:10.18637/jss.v067.i01.

100. Luke SG. Evaluating significance in linear mixed-effects models in R. Behav Res Methods 2017; 49: 1494–1502. DOI: 10.3758/s13428-016-0809-y.

101. Lenth R. Estimated marginal means, aka least-squares means. 2020, p. R package version 1.5.0.

102. Hintze J. PASS 16. Kaysville UT: NCSS, LLC., 2018.

103. Ownby RL, Waldrop-Valverde D, Caballero J, et al. Baseline medication adherence and response to an electronically delivered health literacy intervention targeting adherence. Neurobehav HIV Med 2012; 4: 113–121. DOI: 10.2147/NBHIV.S36549 [doi].

104. Sloan JA and Dueck A. Issues for statisticians in conducting analyses and translating results for quality of life end points in clinical trials. Journal of Biopharmaceutical Statistics 2004; 14: 73–96. DOI: 10.1081/BIP-120028507.

105. Gholami M, Abdoli Talaei A, Tarrahi MJ, et al. The effect of self-management support program on patient activation and inner strength in patients with cardiovascular disease. Patient Educ Couns 2021; 104: 2979–2988. 20210427. DOI: 10.1016/j.pec.2021.04.018.

106. Ownby RL, Waldrop-Valverde D, Jacobs RJ, et al. Cost effectiveness of a computer-delivered intervention to improve HIV medication adherence. BMC Medical Informatics and Decision Making 2013; 13: 29.

107. Gentili A, Failla G, Melnyk A, et al. The cost-effectiveness of digital health interventions: A systematic review of the literature. Frontiers in Public Health 2022; 10. Systematic Review. DOI: 10.3389/fpubh.2022.787135.

108. Moreno R and Mayer RE. Cognitive principles of multimedia learning: The role of modality and contiguity. Journal of Educational Psychology 1999; 91: 358–368. DOI: https://doi.org/10.1037/0022-0663.91.2.358.

109. Klimova B and Zamborova K. Use of mobile applications in developing reading comprehension in second language acquisition--a review study. Education Sciences 2020; 10: 391. DOI: 10.3390/educsci10120391.

110. Sawesi S, Rashrash M, Phalakornkule K, et al. The impact of information technology on patient engagement and health behavior change: A systematic review of the literature. JMIR Med Inform 2016; 4: e1. DOI: 10.2196/medinform.4514.

